# Expression and Distribution of Src Kinase Lyn in the Placenta of Patients with Preeclampsia

**DOI:** 10.64898/2026.10.19.26365934

**Authors:** Taryn D. Heyman, Irina A. Buhimschi, Guomao Zhao, Bani Medegan Fagla, Catalin S. Buhimschi, William E. Ackerman

## Abstract

Src family kinases (SFKs) constitute a major non-receptor tyrosine kinase class that regulate processes important for pregnancy. We profiled placental SFKs to assess cell-specific expression, tissue localization, and clinical associations in early-onset preeclampsia (PE). Placental SFK transcripts were analyzed using bulk RNA sequencing discovery (GSE203507; n=15 PE, n=15 controls) and external validation (GSE114691; n=40 PE, n=21 controls) cohorts. Single-cell and single-nucleus RNA sequencing atlases mapped cell-specific expression. Computational deconvolution and causal mediation analyses were applied to bulk datasets. Findings were cross-validated by real-time quantitative PCR (qPCR; n=9 early-onset PE, n=7 controls) and immunohistochemistry (IHC). Transcriptomic data showed that *LYN* was the sole SFK member consistently elevated in early-onset PE placentas across discovery (1.8-fold, FDR<0.01) and validation cohorts (2.2-fold, FDR<0.001). Elevation was verified by qPCR (p<0.0001) and remained significant after adjusting for clinical covariates. Placental *LYN* abundance correlated positively with maximum antepartum maternal systolic blood pressure (r=0.63, p<0.01). Single-cell transcriptome atlases demonstrated *LYN* enrichment in myeloid lineages, extravillous trophoblasts, and syncytiotrophoblast (SCT). Deconvolution and mediation analyses indicated that altered tissue composition and increased per-cell expression each accounted for approximately half of the PE-associated *LYN* increase. IHC localized Lyn protein to Hofbauer macrophages and the apical microvillar SCT membrane in PE placentas. We conclude that placental *LYN* expression is selectively elevated in early-onset PE and correlates with maternal blood pressure severity. Lyn localization to trophoblasts and placental immune cells suggests potential roles in mediating inflammation and hypoxia-associated signaling in PE pathogenesis.

## 1. Introduction

Hypertensive disorders of pregnancy are among the leading causes of maternal and perinatal mortality in the United States and worldwide [1]. Preeclampsia (PE) represents one of the more complex and consequential of the hypertensive disorders. Manifesting as a progressive syndrome affecting multiple end organs (including the renal, hepatic, pulmonary, and neurologic systems), PE portends serious ramifications for both maternal and fetal health. Early- and late-onset PE differ in epidemiology, presentation, and associated morbidity. Early-onset PE occurs prior to 34 weeks of gestation and is more frequently associated with severe clinical features and fetal growth restriction [2].

Although PE manifests as a multisystem disorder, its pathophysiology is frequently linked to malplacentation [3]. In healthy pregnancies, uteroplacental perfusion is established by placenta-derived extravillous trophoblasts (EVTs), which invade the uterine decidua and remodel maternal spiral arteries into high-capacitance, low-resistance vessels [4]. This process, involving both interstitial and endovascular EVT populations, is essential for normal placental development [4]. In PE, this remodeling is impaired, and as a consequence, placental perfusion is diminished. This leads to the development of maternal disease and fetal growth restriction.

These placentation-associated events are regulated by intracellular signaling pathways, including those mediated by tyrosine kinases, which play key roles in cell growth, lineage specification, and metabolism [5]. Tyrosine kinases are canonically classified into receptor-bound and non-receptor-associated forms [6]. Much of the PE research to date has focused on receptor tyrosine kinases, most notably vascular endothelial growth factor receptor 1 (VEGFR-1) and its isoform, soluble fms-like tyrosine kinase 1 (sFlt-1). Non-receptor tyrosine kinases, in contrast, remain comparatively underexplored.

Src family kinases (SFKs) represent a major class of the non-receptor tyrosine kinases that function within the intracellular compartment to phosphorylate downstream target proteins. SFKs consist of nine members: Src, Fyn, Fgr, Lck, Lyn, Hck, Blk, Yes, and Yrk [7]. Most cells express a variety of SFKs. While some SFKs are broadly expressed (e.g., Src, Fyn, Yes), others exhibit restricted expression patterns (e.g., Lck, Lyn, Hck). Because of their essential functions in numerous cell types, dysregulated SFK signaling is implicated in a variety of disease conditions [8]. Several SFKs function as proto-oncogenes, and there is substantive evidence linking them to the development of solid cancers as well as leukemia via growth factor signaling cascades [9]. Additionally, the role of SFKs in autoimmune and neurodegenerative diseases has been well studied [8]. Term human villous trophoblasts express eight SFK members that follow two transcriptional patterns during cytotrophoblast syncytialization *in vitro*: *FYN*, *HCK*, and *LYN* remain stable, whereas *FGR*, *LCK*, *SRC*, and *YES* rise over a 4-day culture period; *BLK* was detected but could not be followed over time [5]. However, little is known about their involvement in trophoblast differentiation and function during early-onset PE.

As SFK-regulated processes are also important for pregnancy, we set out to determine the relative expression of SFK members in the placenta to explore their potential role in PE. Given that placental hypoxia leads to cellular dysfunction in PE, and SFKs regulate hypoxic stress responses [7], we hypothesized that aberrant SFK expression associates with placental trophoblast and immune dysregulation in PE.

## 2. Materials and Methods

### 2.1 Study Approval

Study approval was granted by the Institutional Review Boards (IRBs) of Yale University, The Ohio State University, the Abigail Wexner Research Institute at Nationwide Children’s Hospital, and the University of Illinois Chicago. All participants provided written informed consent. The currently active IRB approval numbers are STUDY2019-0603 and STUDY2019-0726. For collection ethics pertaining to the secondary analysis of the de-identified public data we defer to the data sources.

### 2.2 Study Population, Materials, and Datasets

To examine placental SFK transcriptional expression profiles, we surveyed our previously reported bulk RNA sequencing dataset GSE203507 [10]. This discovery dataset, cross-sectional in design, included placental villous tissue samples from the following clinical categories: early-onset PE in the absence (n=8) or presence (n=7) of fetal growth restriction (FGR), idiopathic FGR in the absence of hypertension (n=5), spontaneous idiopathic preterm birth controls (sPTB) in the absence of hypertension (n=5), and uncomplicated term deliveries (n=5). These were divided into samples from pregnancies complicated by preterm PE (n=15, gestational age 30±3 weeks [mean±SD]) and normotensive control pregnancies (n=15, gestational age 33±5 weeks).

For independent verification, the RNA sequencing findings from the discovery cohort were cross-referenced with a publicly available, external dataset, GSE114691 (n=61; 21 sPTB controls, 40 early-onset PE cases) [11]. GSE114691 was established to identify differentially expressed genes in placental samples from PE in the absence or presence of FGR relative to normotensive FGR and gestational-age-matched normotensive PTB.

Additional RNA sequencing datasets were used to localize gene expression within placental cell populations. To this end, we used the single-cell and single-nucleus datasets of Admati et al. [12] and the single-cell dataset of Paik et al. (GSE290578) [13]. The Admati et al. [12] datasets were obtained as gene-by-cell unique molecular identifier (UMI) count matrices (depth-normalized) comprising 4 clinical groups: early-onset PE cases (n=10; 32±4 weeks of gestation), idiopathic sPTB controls (n=3; 31±4 weeks), late-onset PE cases (n=7; 38±2 weeks), and term controls (n=6; 39±1 weeks). The GSE290578 dataset comprised 16 libraries from 8 subjects, including early-onset PE (n=4; 31±3 weeks) and term controls (n=4; 39±1 weeks) samples; our analysis was restricted to the deep placenta libraries (GSM8817231, GSM8817233, GSM8817235, GSM8817237, GSM8817239, GSM8817241, GSM8817243, and GSM8817245).

Validation studies were performed using an expanded set of villous placental tissues. For quantitative PCR (qPCR) analysis, the specimens included early-onset PE with severe clinical features (n=9, gestational age 29±1 weeks) and sPTB (n=14, gestational age 29±1 weeks); 9 of these samples were from participants included in the GSE203507 dataset, and 7 were additional specimens selected for this study. For immunohistochemical analysis, we examined early-onset PE (n=14, gestational age 31±3 weeks) and sPTB (n=8, gestational age 31±1 weeks); 7 of these cases were from participants included in the GSE203507 dataset, and 15 were additional cases meeting study criteria. These placental specimens were obtained from our Perinatal Research Biorepository at the University of Illinois Chicago.

### 2.3 Inclusion and Exclusion Criteria

For the GSE203507 dataset and additional samples included in the validation studies, the diagnosis of PE was based on the most up-to-date clinical definition at the time of recruitment [14,15]. Additional details are available in Supplementary Materials.

### 2.4 RNA Sequencing and Data Analysis

The RNA extraction, library preparation, sequencing, and quality control steps were performed as we have previously reported [10]. An independent placenta-derived RNA sequencing dataset, GSE114691 was used to assess the consistency of our findings [11].

The raw FASTQ reads from GSE203507 and GSE114691 were analyzed using our previously described workflow [10,16]. In brief, we first assessed read quality and adapter content with FastQC (v0.11.8). This was followed by adapter removal with Trimmomatic (v0.39). Genome alignment (hg38) and feature count generation used TopHat2 (v2.0.14) and HTSeq (v0.11.2), respectively. The resulting count matrices were screened with the goodSamplesGenes function of WGCNA (v1.72-1), then differential analysis was performed as described below.

For the GSE290578 single-cell dataset, we imported raw matrices using SingleCellExperiment (v1.32.0), then performed quality control (scuttle v1.20.0), doublet detection (scDblFinder v1.24.0), normalization (scran v1.38.1), integration (harmony v2.0.5), and dimensionality reduction (scater v1.38.1). Clustering was performed with Louvain graph partitioning (bluster v1.20.0, k=4, resolution=8), and cellular annotation used CellTypist (v1.7.1) with the Human_Placenta_Decidua model [17]. For the Admati et al. datasets we used the summary files and annotations made publicly available by the authors [12].

For computational deconvolution of the two bulk cohorts (GSE203507 and GSE114691), cell type fractions (θ) were estimated with BayesPrism (v.2.2.3) applying the native Gibbs sampler. The two reference single-cell datasets applied were the Admati and GSE290578 atlases described previously. The posterior coefficients of variation were propagated during downstream analyses. Compositional differences between groups and per-cell expression differences were determined as described in the statistics section.

### 2.5 Real Time qPCR

We used qPCR to validate observations made by RNA sequencing. RNA was extracted and reverse-transcribed to cDNA using random hexamers as we have described previously [18]. Additional details are provided in Supplementary Materials.

### 2.6 Immunohistochemistry

Placental immunohistochemistry (IHC) was conducted using paraffin-embedded tissue sections (∼5 μm). Following deparaffinization and rehydration through graded ethanol, placental sections underwent citrate antigen retrieval (pH 6) and blocking with 5% donkey serum. Sections were incubated overnight at 4°C with primary antibodies against Lyn (1:200; sc-7274, Santa Cruz Biotechnology), CD163 (1:50; MS-1103 Thermo Scientific), or cytokeratin 7 (1:100; 8-0234, Invitrogen), followed by biotinylated donkey anti-mouse IgG (1:600), VECTASTAIN ABC/NovaRed detection (Vector Laboratories), and hematoxylin counterstaining. Normal rabbit IgG served as a negative control.

### 2.7 Statistics

For continuous data, normality was evaluated using the Shapiro-Wilk test. Normally distributed variables were summarized as mean ± standard deviation (SD), otherwise median and interquartile range (IQR; 25th-75th percentiles) were used. Absolute counts and percentages were reported for categorical variables. Point estimates have been reported with 95% confidence intervals (95% CI) for frequentist approaches; 95% credible intervals (95% CrI) were calculated for Bayesian approaches. Independent groups of normally distributed variables were compared using the two-tailed Student t-test; otherwise the Mann-Whitney U test was employed. Pearson correlation analyses were performed for continuous variables. Multivariable linear regression models were fitted using the lm function from the R stats package to evaluate the association between qPCR-measured *LYN* expression and PE, adjusting for covariates (gestational age, birthweight, assigned fetal sex, and delivery mode).

Differential abundance analysis for the bulk RNA sequencing data used DESeq2 (v1.48.2) with design formulas that included clinical group and assigned fetal sex. Pairwise comparisons were evaluated using the Wald test, with multiple comparisons adjustment via the Benjamini-Hochberg false discovery rate (FDR) control procedure. For the single-cell RNA sequencing data, per cell differential expression followed Admati et al. [12] using the Mann-Whitney U test with Benjamini-Hochberg FDR applied to counts normalized to 10,000 UMI per cell.

To test for differences in the deconvolved, four-class cell compositional proportions between clinical groups within each cohort, we performed Dirichet regression using DirichletReg (v0.7-2): a full model predicting class composition by clinical group was compared against an intercept-only null model using a likelihood-ratio test. We additionally assessed these compositional differences using permutational multivariate analysis of variance (PERMANOVA) on sample-level Aitchison distances using the adonis2 function in vegan v.2.7-5 (9,999 permutations).

A mediation analysis was performed by g-formula causal mediation in CMAverse (v0.1.0) to estimate causal randomized-intervention analogs of the natural direct and indirect (i.e., mediated through changes in estimated tissue composition) effects of PE on *LYN* transcriptional abundance through direct counterfactual imputation, following the general procedure described by Campbell et al. [19]. Models were specified using the first 3 principal components of the deconvolved 4-class cell type fractions (θ) vector as simultaneous linear mediators; PE status was the binary exposure, variance stabilizing transformed *LYN* abundance was the outcome, and fetal sex was included as a covariable. Independently, counterfactual decomposition by means of quantile regression (implemented with Counterfactual v1.2) was performed as a second estimate of the extent to which PE-associated shifts in *LYN* abundance were attributable to tissue-level compositional shifts (represented as the isometric log-ratio-transformed deconvolved trophoblast ratio) versus structural expression changes. Effects were evaluated across 9 quantiles using 500 bootstrap iterations.

All statistical tests were two-tailed with significance defined as p<0.05 (with adjustments as specified) or FDR<0.05. GraphPad Prism (v9.3.1; GraphPad Software), SigmaPlot (v14.5; Systat Software), and R (≥v4.4.3; R Foundation for Statistical Computing) were used for statistical analyses.

## 3. Results

### 3.1 Placental LYN Is Elevated in Early-Onset Preeclampsia and Correlates with Maternal Blood Pressure

We first examined the transcriptional levels of Src family kinases in placentas in the absence or presence of early-onset PE using our previously reported bulk RNA sequencing discovery dataset (GSE203507). Among the SFKs, *LYN* was the only family member significantly elevated in participants with early-onset PE compared with normotensive controls (1.8-fold increase, FDR<0.01, Wald test) (Fig. 1A). Using an independent dataset with a larger cohort of placental samples (GSE114691) (Fig. 1B), we verified significant elevation of placental *LYN* in the setting of early-onset PE (2.2-fold increase; FDR<0.001, Wald test). In addition to *LYN*, analysis of this second dataset also revealed a modest elevation in *YES1* (1.1-fold; FDR<0.05) and significant decreases in *HCK*, *BLK*, and *FYN* in PE (0.6-fold, 0.4-fold, and 0.9-fold, respectively; FDR<0.01).

**Fig. 1.**
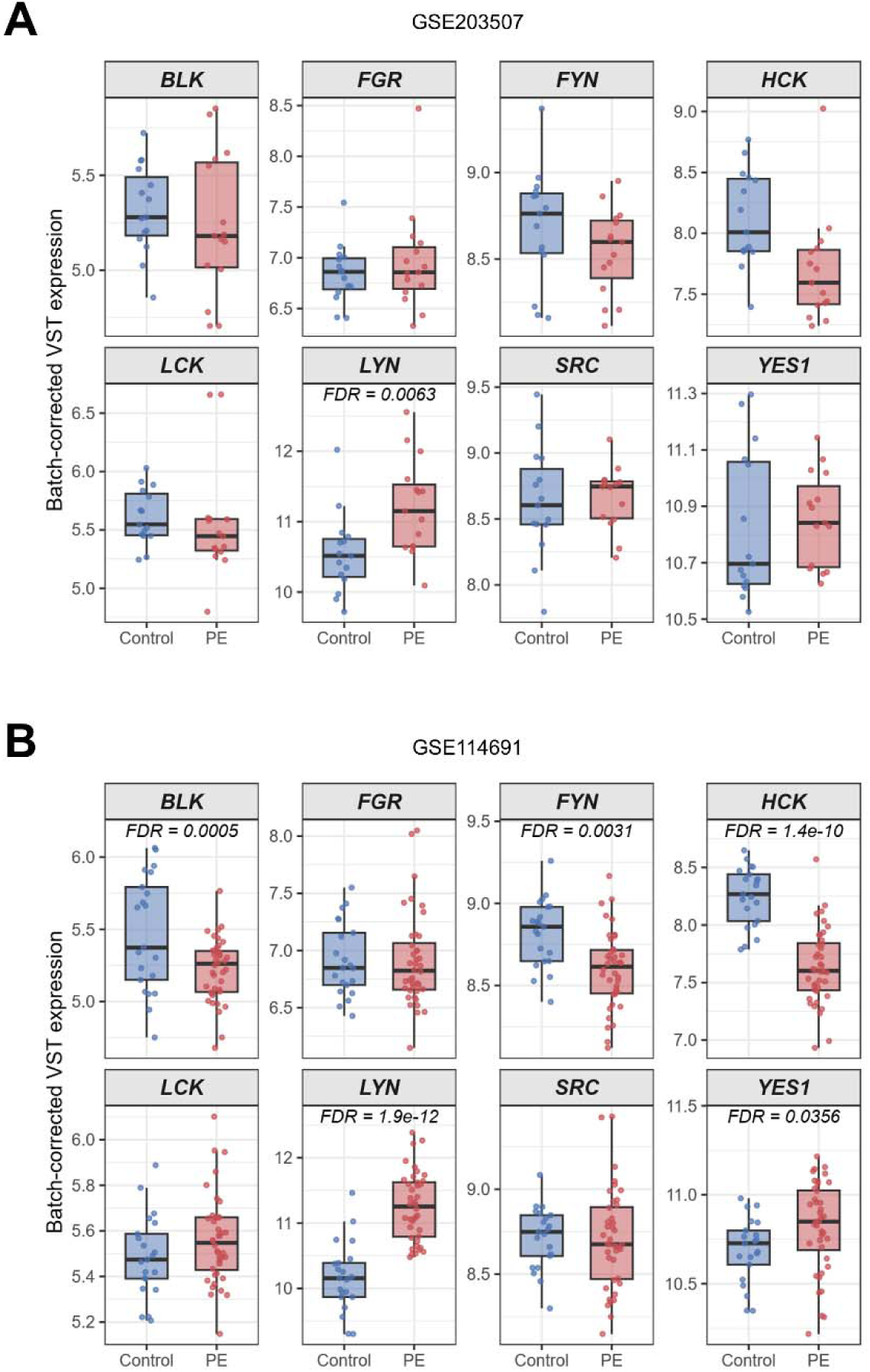
Placental Src family kinase (SFK) transcriptional expression in early-onset preeclampsia. Box plots with overlaid data points showing batch-corrected variance-stabilized (VST) expression for the eight human SFKs (*BLK*, *FGR*, *FYN*, *HCK*, *LCK*, *LYN*, *SRC*, *YES1*) in (A) GSE203507 (n=15 control, n=15 PE) and (B) GSE114691 (n=21 control, n=40 PE). Boxes span interquartile range, horizontal line denotes median. Differential abundance between pairs was tested with the Wald test in the DESeq2 (v1.48.2) workflow.

To cross-validate these findings, qPCR was performed using an expanded validation cohort (n=7 sPTB; n=9 early-onset PE). We verified that placental *LYN* mRNA was significantly elevated in preterm PE compared to sPTB controls (p<0.0001, t-test). These associations remained significant when stratified into the original (n=3 sPTB, n=6 PE; p<0.05) and newly added (n=4 sPTB, n=3 PE; p<0.001) sample sets, as well as in multivariable models following adjustment for gestational age, birthweight, fetal sex, and mode of delivery (p<0.001) (Fig. 2A).

**Fig. 2.**
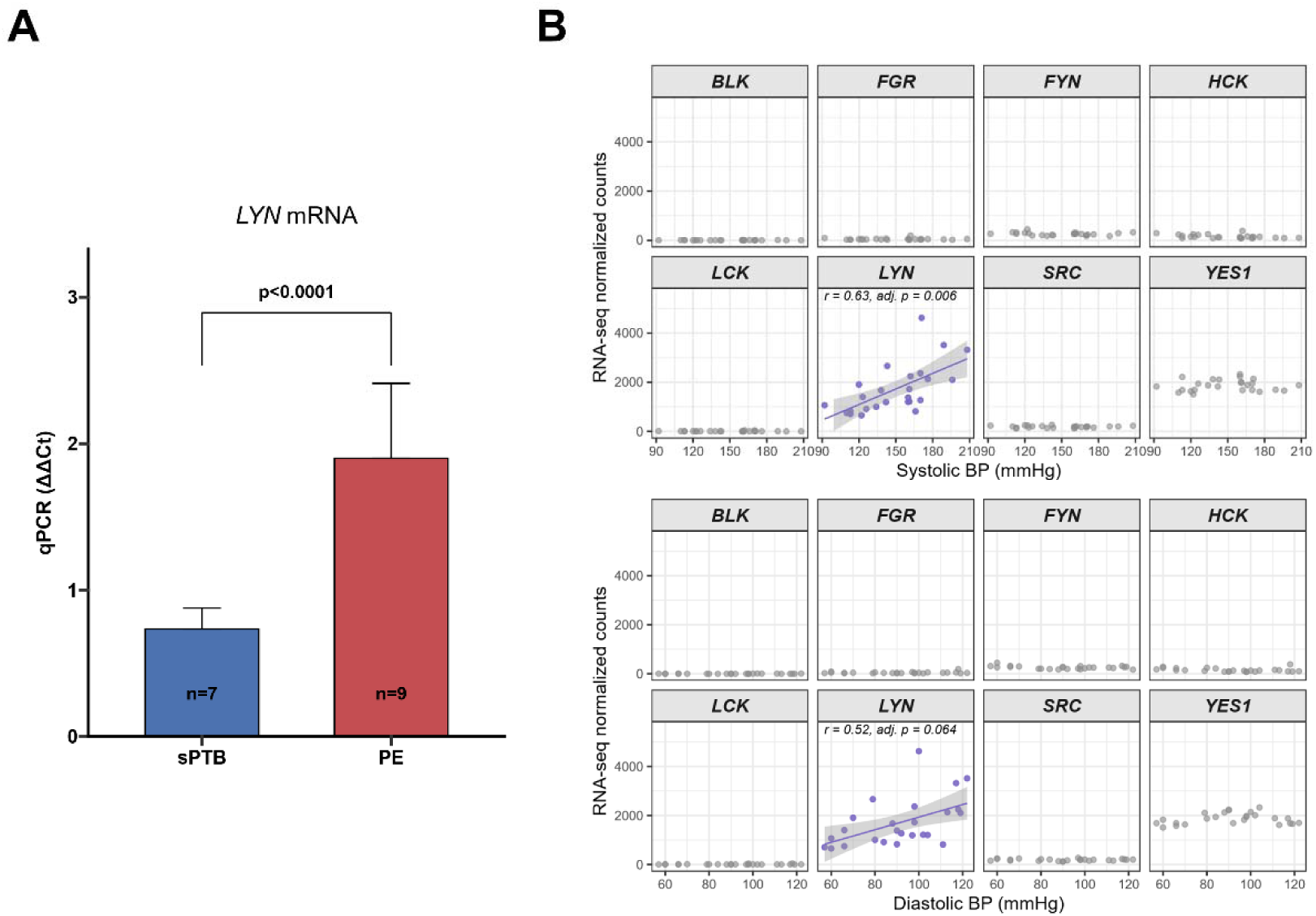
Placental *LYN* transcriptional abundance is elevated in early-onset preeclampsia and correlates with maternal systolic blood pressure. (A) Relative placental *LYN* transcript levels were quantified by qPCR in an expanded validation cohort of spontaneous preterm birth controls (sPTB, n=7) and early-onset preeclampsia (PE, n=9) placenta specimens. Bars represent mean ± SD. Following Shapiro-Wilk testing, differences in ΔΔCt were tested using the unpaired two-tailed Student t-test with Welch correction; t(9.55)=6.52, p<0.0001. The difference persisted when the cohort was partitioned into the original (n=3 sPTB, n=6 PE) and newly added (n=4 sPTB, n=3 PE) sample sets analyzed individually, and in multivariable linear models adjusted for gestational age at delivery, birthweight, fetal sex, and mode of delivery. (B) Association between placental Src family kinase expression and maternal blood pressure in GSE203507 (n=25). Normalized feature counts plotted against highest recorded antepartum systolic (top panel) and diastolic (bottom panel) maternal blood pressures. For *LYN* (purple) a linear regression line with 95% confidence interval is overlaid. Correlations are Pearson coefficients with Bonferroni-adjusted p values.

Next, to evaluate whether placental SFK expression correlated with clinical PE features, we examined the relationships with maternal clinical parameters. In our GSE203507 dataset, we found that *LYN*, but not the remaining SFKs, correlated positively with highest recorded antepartum maternal systolic blood pressure (r=0.63; p_adj_<0.01, Bonferroni-corrected) (Fig. 2B). *LYN* was also nominally correlated with diastolic blood pressure (r=0.52; p<0.01), but this relationship failed to reach significance following adjustment for multiple comparisons (p_adj_=0.064, Bonferroni-corrected).

### 3.2 Cell Type-Specific Expression and Placental Localization of Lyn

Insofar as *LYN* was the sole SFK consistently elevated in PE across multiple cohorts, and given its unique correlation with maternal systolic blood pressure, we prioritized it for further investigation.

To identify the placental cell types expressing *LYN* mRNA, we queried the single-cell RNA sequencing dataset published by Admati et al. [12]. This dataset comprised 86,752 cells isolated from placental cotyledons from 26 donors (10 early-onset PE cases, 3 normotensive idiopathic sPTB controls, 7 late-onset PE cases, and 6 term controls). Cells were annotated using the metadata provided by the original authors, who identified 46 cell types in 6 general classes (Fig. 3A). Within this compendium, *LYN* was enriched in myeloid, stromal, and lymphoid cells, and found in lower proportions among trophoblastic, endothelial, and mural cells (Fig. 3B,C).

**Fig. 3.**
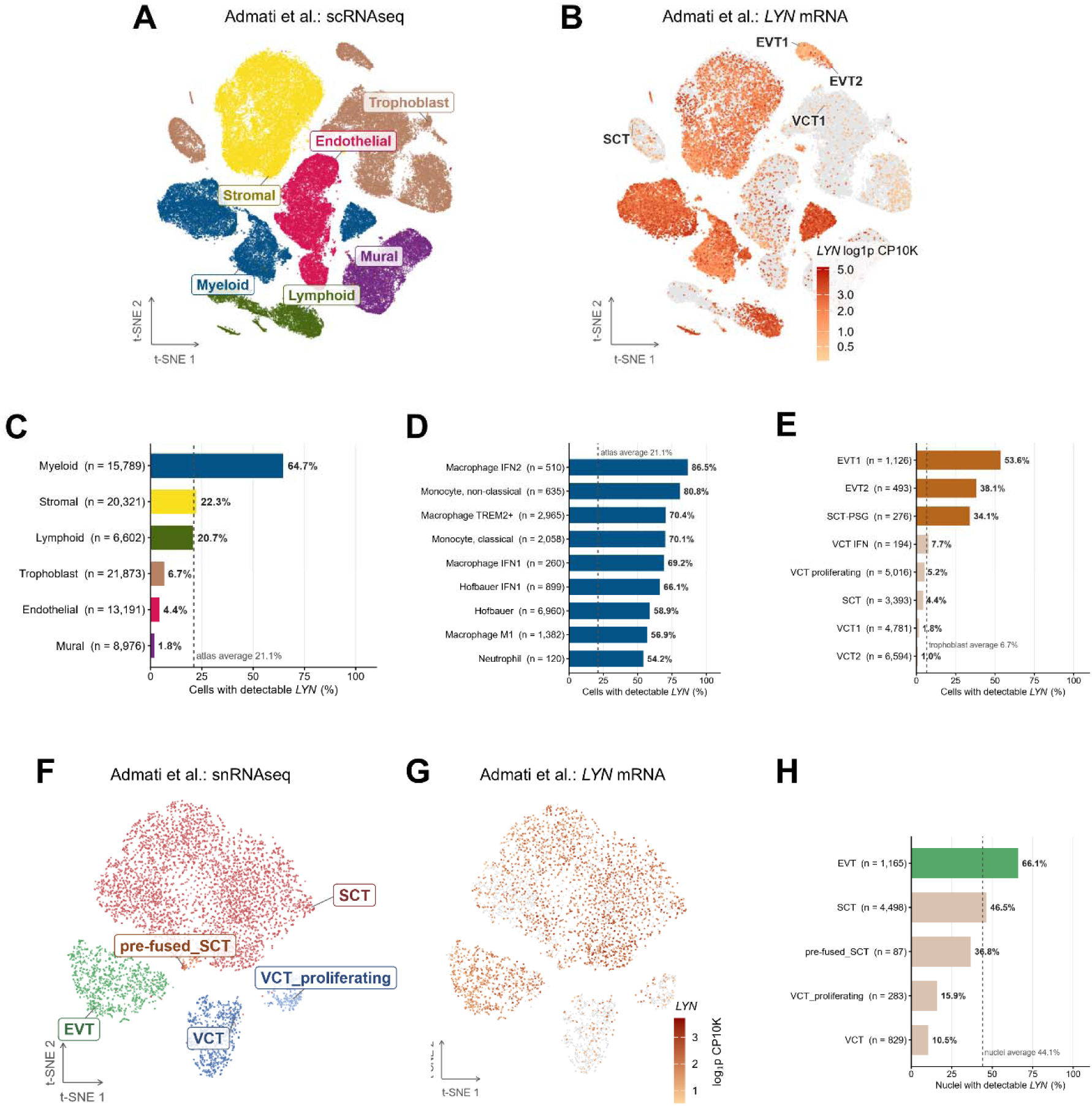
*LYN* transcript detection in individual placenta cell populations and nuclei. (A) Reanalysis of 86,752 individual placental cells from the Admati et al. dataset visualized using a t-distributed stochastic neighbor embedding (t-SNE) plot. (B) *LYN* expression in the t-SNE plot as indicated by orange color saturation; 18,312 (21.1%) of placental cells had detectable *LYN*. Units are log-transformed counts (offset by 1) per 10,000 transcripts (log1p CP10K). (C) Percentages of cells per lineage group with detectable *LYN*. (D) Percentages of myeloid sublineages with detectable *LYN*. (E) Percentages of trophoblast sublineages with detectable *LYN*. (F) t-SNE plot showing 6,862 single nuclei from the Admati et al. dataset colored by major cell type. (G) *LYN* expression in the t-SNE plot as indicated by brown color saturation, expressed as log1p CP10K; 3,024 (44.1%) of trophoblast nuclei had detectable *LYN*. (H) Percentages of nuclei with detectable *LYN* among the trophoblast sublineages. Reported detection rates may underestimate true detection in deeply sequenced cells as low counts round to zero after normalization. Abbreviations: EVT, extravillous trophoblast; IFN, interferon; VCT, villous cytotophoblast, scRNAseq, single-cell RNA sequencing; SCT, syncytiotrophoblast; snRNAseq, single-nucleus RNA sequencing; TREM, triggering receptor expressed on myeloid cells.

Among the molecularly distinct myeloid sublineages, *LYN* detection was highest in interferon (IFN)-activated macrophages (86.5%) and non-classical monocytes (80.8%), and exceeded 54% in the remaining subtypes, including placental macrophages (Hofbauer cells) and neutrophils (Fig. 3D). In the trophoblastic lineage, *LYN* mRNA detection was largely confined to EVT molecular subtypes (EVT1 53.6%, EVT2 38.1%) and the pregnancy-specific beta-1 glycoprotein-expressing (PSG) syncytiotrophoblast (SCT) subtype (34.1%); *LYN* detection was low in the villous cytotrophoblast (VCT) subtypes (VCT1 1.8%, VCT2 1.0%, proliferating VCT 5.2%) and bulk SCT (4.4%) (Fig. 3E). To corroborate these findings, we additionally interrogated a second placental single-cell dataset [13] (GSE290578; 4 term samples, 4 early-onset PE samples). Insofar as this study concerns placental *LYN*, we used only the deep placenta (villous tissue) library from each subject. Across 58,900 cells that resolved into 19 cell types, the myeloid lineage again showed the highest proportion of detectable *LYN* expression (67.6% with detectable *LYN*), whereas in trophoblasts (30.5% with detectable *LYN*), *LYN* displayed ∼12-fold enrichment in EVT compared to VCT (56.0% vs. 4.7%). (Supplemental Fig. 1)

As a multinucleated syncytium, SCT is inefficiently recovered using dissociation-based single-cell methods; therefore we also assessed *LYN* mRNA expression in the single-nucleus RNA sequencing dataset generated by Admati and colleagues [12] (6,862 nuclei from 5 donors comprising 3 early-onset PE and 2 sPTB control samples). The nuclei were annotated by the original authors into five subtypes (SCT, EVT, VCT, proliferating VCT, and pre-fused SCT) (Fig. 3F). Within this atlas, which contained a higher proportion of syncytium-derived material than the companion single-cell dataset, *LYN* mRNA was detected in 66.1% of EVT, 46.5% of SCT, 36.8% of pre-fused SCT, 15.9% proliferating VCT, and 10.5% of VCT (Fig. 3G,H).

To validate these findings at the protein level, we next performed IHC analysis (Fig. 4). In placental sections from sPTB controls, Lyn-like immunoreactivity was most pronounced within villous stromal cells exhibiting ovoid and stellate-to-dendritic morphologies (Fig. 4A). In PE, in addition to villous stromal cells, anti-Lyn labeled the syncytiotrophoblast along the apical and basal membranes (Fig. 4B,C). Hypothesizing that the observed stromal immunolabeling represented Hofbauer cells, we examined serial placental sections labeled with antibodies directed against Lyn, CD163 (a macrophage marker), and cytokeratin 7 (CK7, a cytotrophoblast marker). The serial sections demonstrated that the villous core Lyn labeling pattern was consistent with CD163^+^ Hofbauer staining (Fig. 4D,E), while prominent trophoblast labeling was appreciated in the apical microvillous membrane (Fig. 4D,F).

**Fig. 4.**
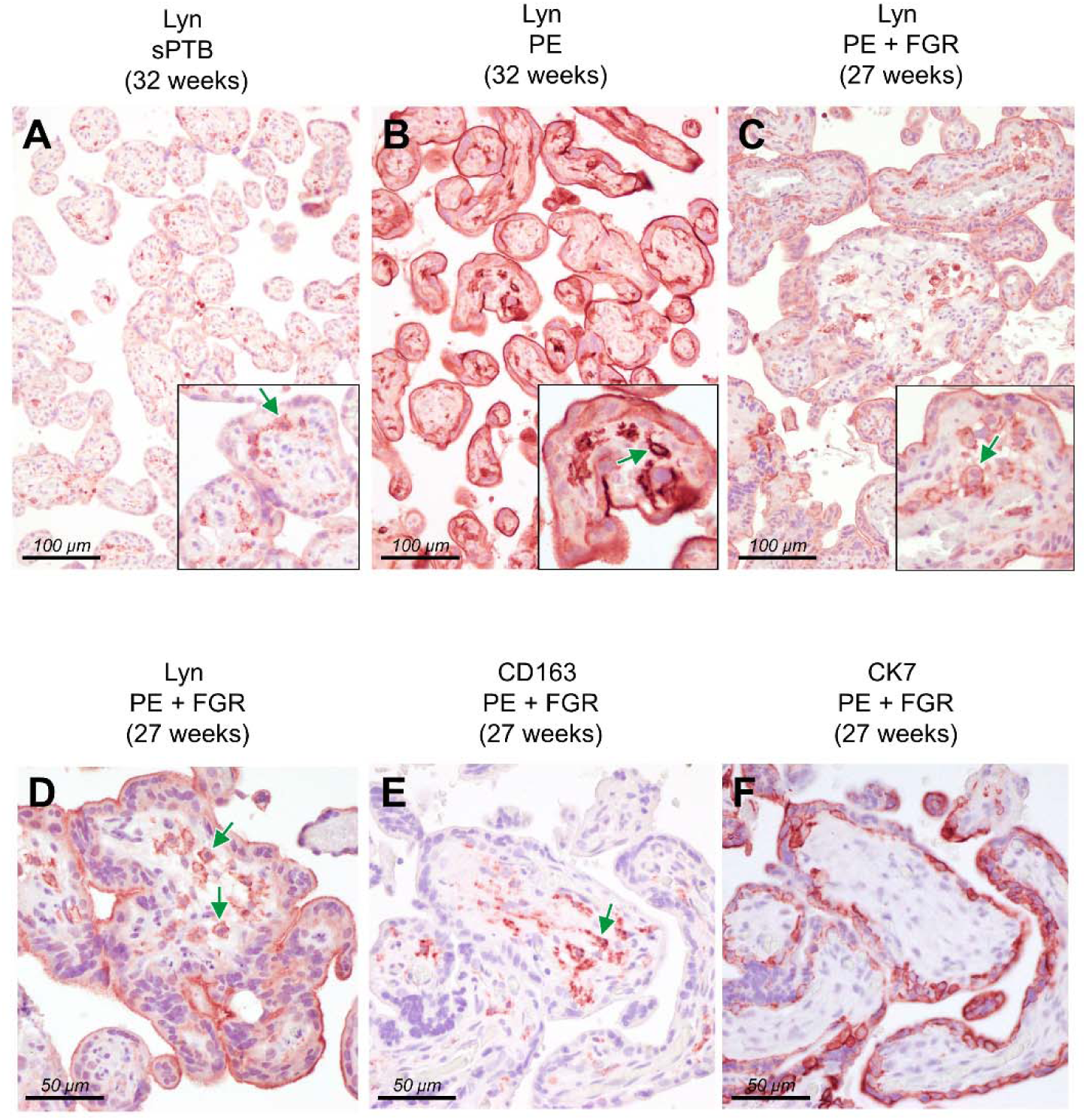
Immunohistochemical localization of Lyn protein in placental specimens. (A-C) Formalin-fixed, paraffin-embedded villous placenta specimens stained with anti-Lyn (1:200). Representative photomicrographs demonstrate Lyn-like immunoreactivity (red) in preterm placental specimens with gestational ages at delivery indicated. (A) Lyn localization in normotensive spontaneous preterm birth (sPTB, delivered at 32 weeks of gestation). Inset shows detail of labeling within villous stromal cells (green arrow). (B) Lyn localization within the syncytiotrophoblast apical and basal membranes, in addition to villous stromal cells (inset, green arrow), in the setting of early-onset preeclampsia (PE, 32 weeks). (C) Lyn localization in the setting of PE complicated by fetal growth restriction (FGR) at 27 weeks of gestation, with inset showing villous stromal cell labeling (green arrow). (D-F) Serial sections of specimens (PE + FGR) were stained with anti-Lyn (D), anti-CD163 (1:50) to identify Hofbauer cells (placental macrophages) (E), and anti-CK7 (1:100) to identify trophoblasts (F). Green arrows in panels D and E show that the observed villous stromal labeling is consistent with Hofbauer cells. Scale bars, 100 μm (top row), 50 μm (bottom row).

### 3.3 Cellular deconvolution reveals altered placental composition and increased per-cell LYN ***expression in early-onset preeclampsia***

Bulk RNA sequencing cannot distinguish between elevated *LYN* expression within individual cells and an expansion of *LYN*-expressing cellular populations within samples. Prior reports have indicated that a proportion of the transcriptional abundance changes in bulk sequencing data may arise from altered placental cellular composition in PE [19,20]. To resolve these ambiguities, we utilized the single-cell reference atlases to computationally deconvolve cell type proportions within the bulk datasets. The capture profile of the Admati et al. [12] single-cell atlas was balanced across cellular compartments (∼25% each for trophoblast, stromal, vascular, and immune cells), while the GSE290578 single-cell dataset was biased toward immune populations (74.5% immune cells captured vs. 12.7% trophoblast, 10.5% stromal, and 2.2% vascular) (Fig. 5A). Deconvolution of the GSE203507 and GSE114691 bulk datasets revealed that most of the total mRNA transcript mass was attributable to trophoblasts (54-72% across both cohorts and clinical groups) (Fig. 5A). The deconvolved cell-type composition in these placental samples differed significantly between the control and early-onset PE groups in both cohorts (p<0.001 for each within-cohort comparison, Dirichlet likelihood-ratio test; p<0.005 for each within-cohort comparison, PERMANOVA). In both bulk datasets, the trophoblast RNA fraction increased in PE relative to control samples, while that of the stromal, vascular, and immune fractions declined. The bulk *LYN* transcript abundance correlated positively with the deconvolved trophoblast fraction in both datasets (GSE203507: r =0.66, p<0.001; GSE114691: r=0.78, p<0.001; note that the strength and statistical significance of these associations were robust to compositional closure when evaluated using isometric log-ratio transformation of trophoblasts versus remaining cell types). To estimate the extent to which altered cellular composition might account for the observed *LYN* increase, we used two independent approaches: g-formula causal mediation analysis, and counterfactual decomposition regression (Supplemental Fig. 2). In the GSE114691 dataset, these estimators suggested that approximately half of the PE-associated increase in *LYN* abundance could be attributed to altered composition: 57.7% (95% CI: 34.3-86.2%; g-formula mediation), and 56.3% at the median (95% CI: 27.1-85.5%; distributional decomposition). In GSE203507, both methods returned point estimates of similar magnitude: 39.2% (95% CI: −12.9-120.6%; g-formula mediation), and 67.5% at the median (95% CI: 14.2-120.8%; distributional decomposition).

**Fig. 5.**
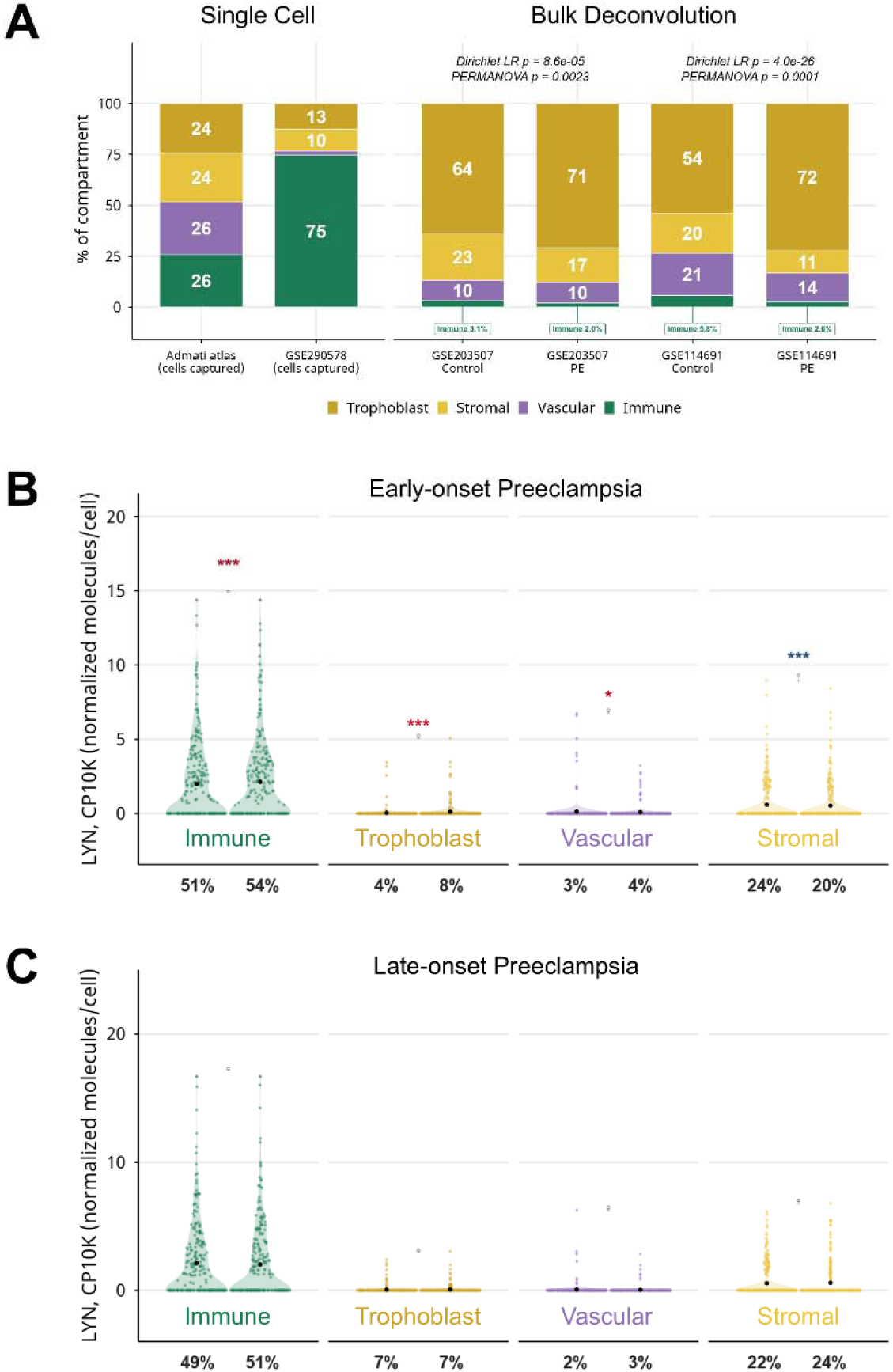
Placental cellular composition and lineage-specific *LYN* transcriptional expression in preeclampsia. (A) Cellular composition in single-cell protocol capture profiles (Admati et al. dataset and GSE290578) and deconvolved bulk RNA sequencing data (GSE203507 and GSE114691). On the left, stacked bar charts display the percentages attributed to each major cell class in each single cell dataset. On the right, the proportions of deconvolved mRNA transcript mass attributed to each major cell class (as estimated using BayesPrism v.2.2.3 with the Admati atlas as reference) are shown. For the deconvolved proportions, the credible intervals were all within ±0.017 percentage points of the target point estimates. Compositional changes between normotensive Control and early-onset preeclampsia (PE) groups were tested in each cohort using a Dirichlet regression likelihood-ratio test (Dirichlet LR), then cross-checked with a PERMANOVA on Aitchison distances. Labels inside bars represent mean percentages per compartment; immune cell fractions (<6%) are annotated with leader lines. (B) Sina plots showing normalized *LYN* counts (per 10,000 transcripts; CP10K) by lineage class for early-onset preeclampsia cases in the Admati et al. dataset. Percentages under the axis are the detection rate (percent of cells with ≥1 detected *LYN* transcript). (C) Sina plots as in panel B showing normalized per-cell *LYN* mRNA expression in late-onset PE cases in the Admati atlas. Colored asterisks (red, increased in PE; blue, decreased in PE) represent two-sided Wilcoxon rank-sum tests on log(1 + CP10K), adjusted using the Benjamini-Hochberg procedure. Black dots in panels B and C represent mean values.

Next, to address whether per-cell *LYN* expression changed within individual cell populations, we probed the Admati et al. [12] single-cell dataset, in which the confounding effects of gestational age were controlled for by study design. This analysis demonstrated increased *LYN* expression in immune, trophoblast, and vascular cells in the early-onset PE group (compared to gestational age-matched normotensive controls) and decreased *LYN* in the stromal cell populations (FDR<0.001 for immune, trophoblast, and stromal subpopulations; FDR<0.05 for vascular; Mann-Whitney U test). (Fig. 5B). In contrast, none of these compartments showed significant differences in per-cell *LYN* expression between late-onset PE and term controls (all FDR>0.05) (Fig. 5C). At finer resolution, the per-cell increase in *LYN* was unevenly distributed across the molecularly-defined sublineages (Supplemental Fig. 3). Significant differences (FDR<0.05 within the 44-cell-type family; Mann-Whitney U test with Benjamini-Hochberg correction) were confined to natural killer (NK) lymphocytes and proliferating cells in the immune compartment, SCT within the trophoblast population, and the GALNT15^+^ subtype within the stromal compartment. These latter results should be interpreted cautiously, however, as sparse *LYN* detection within some sublineages (in which the differential analysis rests on a smaller absolute number of *LYN*-expressing cells) limits the ability to draw robust inferences.

## 4. Discussion

### Principal findings

In this report, we tested the hypothesis that altered SFK expression associates with PE. Using two independent bulk RNA sequencing cohorts, in addition to a target-validating qPCR cohort, we established that among the SFKs, *LYN* mRNA was selectively elevated in early-onset PE placentas compared to normotensive controls. This elevation persisted after controlling for clinical confounders (including gestational age, birthweight, fetal sex, and mode of delivery) and correlated positively with maximum antepartum maternal systolic blood pressure. Furthermore, statistical modeling approaches demonstrated that increased *LYN* in early-onset PE placentas reflected changes in tissue composition as well as altered cellular expression, with each accounting for approximately half of the overall observed effect.

At the cellular level, by integrating single-cell and single-nucleus transcriptomic datasets with IHC, we found that placental *LYN* mRNA was overrepresented in immune cells (particularly myeloid lineages) and stromal cells. Within the trophoblast lineage, *LYN* expression was enriched in EVTs and SCT. Protein-level validation confirmed Lyn localization to CD163^+^ Hofbauer cells, with accumulation along the apical microvillous membrane of the SCT (hemochorial interface) in PE tissues.

### Results in the context of what is known

Lyn, a member of the Src family of non-receptor tyrosine kinases, differs fundamentally from the receptor tyrosine kinases that have received significant attention in PE research [21]. Whereas receptor tyrosine kinases such as VEGFR1 directly bind extracellular ligands and transduce signals across the plasma membrane, Lyn lacks an extracellular ligand-binding domain and instead functions intracellularly as a signaling intermediary downstream of multiple cell-surface receptors [22]. Through phosphorylation of downstream substrates, Lyn can integrate signals arising from growth factor, immune, adhesion, and inflammatory pathways and thereby influence cellular proliferation, migration, differentiation, cytoskeletal organization, and inflammatory responses [23].

Lyn expression is well-documented in macrophages, B lymphocytes, and vascular endothelial cells [24,25]. In addition to detecting Lyn in myeloid, stromal, lymphoid, and endothelial cells, we report its presence within placental trophoblasts, with marked enrichment in two fulcral lineages: (1) EVTs, which invade the uterus to remodel maternal spiral arteries and establish low resistance channels that irrigate the intervillous space; and (2) SCT, the hemochorial barrier directing selective maternal-fetal exchange. Our results corroborate and extend those of Daoud et al. [5], who reported the full complement of SFK expression (including two alternatively spliced Lyn isoforms) in primary villous trophoblasts isolated from human term placenta.

In early-onset PE, elevated *LYN* in bulk placental RNA-sequencing data reflects, in part, altered tissue composition. Using reference-based cell-type deconvolution, Campbell et al. [19] demonstrated that PE placentas harbor an increased proportion of EVTs, consonant with maturation arrest, aberrant interstitial invasion, and the impaired adhesion-phenotype switch originally characterized by Zhou and colleagues [26,27]. *LYN* mRNA enrichment in EVTs accords with recent work by Kumar et al. [28] and suggests a role in migration and invasion, both essential underpinnings for successful implantation. This is plausible given that SFKs regulate focal adhesion turnover and matrix metalloproteinase expression in other migratory cell types [21,29–31].

In the SCT, Lyn likely serves multifarious roles. Lyn is upregulated at the trophoblast plasma membrane during syncytialization and promotes efficient CTB fusion *in vitro* [32]. Beyond this role in syncytialization, it is tenable that Lyn enrichment at the SCT apical surface supports plasma membrane repair (e.g., during SCT shedding or in response to mechanical or ischemic strain). Trophoblasts (including the SCT) reseal focal membrane disruptions through an annexin-dependent mechanism [33] which, in other cell types, requires SFK activity [34]. SFKs also regulate signaling through Siglec-6 (sialic acid-binding immunoglobulin-like lectin-6) [35], which is expressed in SCT and increased in PE [36]. This pathway is causally linked to PE-like pathology in mice [37]. Whether Lyn is specifically involved in these processes or related pathways remains to be determined.

Among leukocyte populations, Lyn operates as a modulator of immune tone. In myeloid cells, Lyn generally contributes to lineage-specific functions including survival and proliferation (e.g., transducing growth factor signals in monocytes and macrophages), inflammatory regulation (e.g., dampening mast cell degranulation and tuning cytokine production), and effector responses (e.g., driving antibody-mediated phagocytosis and respiratory burst in neutrophils) [21,30]. Campbell and colleagues [19] reported a decrease in the proportion of Hofbauer macrophages in PE tissue, complementing prior work by Tang et al. [38] and buttressing our current observations. Lyn normally restrains myeloid activation [30], and loss of this inhibitory signal alongside tolerogenic placental macrophages may promote a pro-inflammatory microenvironment. In the single-cell data, we also observed that *LYN* mRNA detection was higher in proliferating leukocytes and NK lymphocyte populations in early-onset PE. Given that SFKs, including Lyn, govern signaling tone for both activating and inhibitory immunoreceptors (such as the killer immunoglobulin-like receptors [KIRs] which engage fetal HLA antigens on EVTs to promote placentation) [24,39], this may indicate recalibration in these populations in response to placental antigens. Since inhibitory maternal-fetal combinations are themselves associated with PE risk [40], it is possible that elevated Lyn in NK cells contributes to dysregulated signaling at decidual NK-EVT synapses, contributing to the apparent stall in EVT invasion described above [41]. We raise these hypotheses as potential directions for future research (see also the research implications section below).

### Clinical implications

*Prima facie*, Lyn provides a compelling pharmaceutical target to address the pathophysiology of PE. As a regulator downstream of multiple signaling effectors, Lyn modulates endothelial cell activation, microvascular permeability, and immune cell responsiveness. Pharmacological inhibition of this pathway could ameliorate systemic maternal endothelial dysfunction, a hallmark of severe PE phenotypes.

Current therapies targeting SFKs are primarily utilized in oncology and rheumatology. The SFK-inhibiting FDA approved drugs dasatinib, bosutinib, and ponatinib are effective in treating chronic myeloid leukemia [42]. Pharmacological drugs targeting these pathways provide indiscriminate kinase coverage rather than isoform selectivity and span the Bcr-Abl oncoprotein tyrosine-kinase inhibitor derivatives (dasatinib, bosutinib), off-target inhibitors (ponatinib, nintedanib), and tirbanibulin (designed to target Src directly and used topically for actinic keratosis). Although Lyn possesses a ligandable ATP-binding pocket, high active-site conservation among SFKs has constrained Lyn-selective inhibition [43].

Systemic SFK inhibitors are generally avoided in pregnancy, however. Tyrosine kinase inhibitors interfere with basic intracellular signaling pathways involved in cellular growth, proliferation, and differentiation. Therefore, exposure to medication inhibiting tyrosine kinases during early organogenesis can lead to congenital defects, spontaneous abortion, and have adverse effects on fetal growth [44].

### Research implications

Identification of Lyn in the placenta expands the range of tyrosine kinase signaling pathways potentially involved in PE beyond the well-established receptor-mediated pathways. The reproducible increase in *LYN* transcript abundance across independent cohorts, its relationship with maternal systolic blood pressure, and its localization to specific placental cell populations provide a rationale for further investigating whether altered Lyn signaling contributes functionally to the PE phenotype.

In early-onset PE, placental malperfusion produces hypoxia-reoxygenation injury and endoplasmic reticulum dysfunction, both of which contribute to oxidative stress, an established, direct Lyn activator [45,46]. Transcript and protein abundances, however, are imperfect proxies for kinase activity, and future work will be required to quantify Lyn phosphorylation states and catalytic activity in these tissues. Additionally, Lyn exists as two protein isoforms that differ by 21-amino-acids in the N-terminal region. LynA, the full length protein, undergoes rapid ubiquitin-mediated degradation upon activation, whereas LynB, lacking the regulatory residues (particularly Y32) that serve as degradation markers, persists throughout the signaling response [47]. Although both are present in placental cells [5], we were unable to reliably resolve isoform use in the available bulk RNA-sequencing datasets due to insufficient read coverage across the alternative exon junction, and additional studies are needed to resolve Lyn isoform expression in the context of PE.

### Strengths and limitations

Strengths of this study include the robust multi-dataset design with cohort replication, utilization of systems-level transcriptomic datasets, multi-platform cross-validation, and the thoroughness in which findings were interrogated using multiple independent approaches.

Limitations include the relatively small sample size in some subgroups, the observational nature, and the cross-sectional design which constrains causal inference. Furthermore, we acknowledge methodological caveats, including the approximate nature of cell-deconvolution estimates, cross-sectional mediation reflecting quantitative decomposition rather than causal sequence, and potential confounding from underlying pathologies in gestational-age-matched preterm controls. This is a descriptive study, and must be complemented with loss- or gain-of-function experiments to evaluate whether Lyn contributes to trophoblast, macrophage, and lymphocyte phenotypes. Finally, our findings in early-onset disease should not be extrapolated to late-onset PE.

### Conclusion

Placental LYN expression is significantly increased in early-onset PE and correlates with maternal blood pressure severity. Increased localization of LYN within SCT, EVTs, and placental immune cells suggests that this Src family kinase may contribute to PE pathobiology through inflammatory and hypoxia-associated signaling mechanisms. These findings identify Lyn as a potential molecular mediator and therapeutic target in PE.

## Statements and Declarations

### Author Contributions

Conceptualization: TDH, IAB, CSB, WEA. Methodology: TDH, IAB, GZ, BMF, CSB, WEA. Formal analysis and investigation: TDH, IAB, GZ, BMF, CSB, WEA. Writing – original draft preparation: TDH, WEA. Writing – review and editing: TDH, IAB, GZ, BMF, CSB, WEA. Resources: IAB, CSB. Supervision: IAB, CSB, WEA. All authors read and approved the final manuscript.

### Ethics Approval

All procedures performed in studies involving human participants were in accordance with the ethical standards of the institutional and/or national research committee and with the 1964 Helsinki Declaration and its later amendments or comparable ethical standards. The study was approved by the Institutional Review Boards of Yale University, The Ohio State University, the Abigail Wexner Research Institute at Nationwide Children’s Hospital, and the University of Illinois Chicago (currently active approvals STUDY2019-0603 and STUDY2019-0726).

Secondary analyses of de-identified public datasets relied on the ethics approvals of the original studies.

### Consent to Participate

Written informed consent was obtained from all individual participants included in the study.

### Data Availability

The bulk RNA sequencing datasets analyzed during the current study are available in the NCBI Gene Expression Omnibus (GEO) under accession numbers GSE203507 and GSE114691, and the single-cell and single-nucleus datasets are available under GEO accession number GSE290578 and Figshare identifiers 23264102, 23628471, and 23264165. Other data supporting the findings of this study are available from the corresponding author on reasonable request, subject to institutional approval.

## Acknowledgments

Portions of this work were presented in abstract form at the 44th Annual Pregnancy Meeting of the Society for Maternal-Fetal Medicine, Feb 10-14, 2024, National Harbor, MD. Grammarly for Windows (version 1.2.292.1950, Grammarly, Inc.), Google Gemini (≥3.6), and Anthropic Claude (Sonnet ≥4.6, Opus ≥4.8) were used for editorial support to improve clarity and grammar during manuscript preparation and revision. For data analysis, the initial R scripts were hand-coded using routine workflows, then Claude AI was used to audit all logic, refactor the analytic pipeline, and perform additional quality assurance checks (reviewer mode). The authors reviewed and edited all content to ensure accuracy and originality. The authors take full responsibility for the content of the final publication, and all intellectual contributions remain our own.

