## Supplemental Material for "Expression and Distribution of Src Kinase Lyn in the Placenta of Patients with Preeclampsia"

**Acknowledgments**

Portions of this work were presented in abstract form at the 44th Annual Pregnancy Meeting of the Society for Maternal-Fetal Medicine, Feb 10-14, 2024, National Harbor, MD. Grammarly for Windows (version 1.2.292.1950, Grammarly, Inc.), Google Gemini (≥3.6), and Anthropic Claude (Sonnet ≥4.6, Opus ≥4.8) were used for editorial support to improve clarity and grammar during manuscript preparation and revision. For data analysis, the initial R scripts were hand-coded using routine workflows, then Claude AI was used to audit all logic, refactor the analytic pipeline, and perform additional quality assurance checks (reviewer mode). The authors reviewed and edited all content to ensure accuracy and originality. The authors take full responsibility for the content of the final publication, and all intellectual contributions remain our own.

**Table of** **Contents**

### S1.1 Supplemental Materials and Methods

### S1.1 Detailed Inclusion and Exclusion Criteria

Severe early-onset preeclampsia (PE) was diagnosed prior to 34 weeks of gestation by one or more of the following: (1) systolic blood pressure of >160 mm Hg or diastolic >110 mm Hg on at least 2 occasions 6 h apart; (2) >5 g protein excretion per 24 h urine collection, and/or persistent +3 proteinuria on dipstick testing; (3) oliguria <500 ml/24 h; (4) cerebral or visual disturbances; (5) pulmonary edema or cyanosis; (6) epigastric or right upper quadrant pain; (7) impaired liver function; and/or (8) thrombocytopenia. Spontaneous preterm birth (sPTB) was defined as the spontaneous labor onset with delivery before 37 gestational weeks. For all cases of sPTB, triple I was excluded (negative amniotic fluid biochemical tests, negative bacterial cultures, and absence of histologic chorioamnionitis or funisitis). Pregnancies with multifetal gestations, fetal structural anomalies, chromosomal aneuploidies, and known comorbidities (e.g., diabetes, thrombophilias, thyroid disease, etc.) were excluded. Gestational age was calculated by last menstrual period and/or ultrasound measurements before 20 weeks of gestation.

### S1.2 Detailed Real Time qPCR

Quantitative real-time PCR was performed using TaqMan reagents (Applied Biosystems, Foster City, CA) in 20-µL reactions comprising 10 µL mastermix (TaqMan Fast Universal PCR 2×Master Mix), 8 µL water, 1 µL cDNA template normalized, and 1 µL PCR primer-probe set (TaqMan Gene Expression Assay hs00199831_m1). For endogenous controls, we used a pair of TaqMan primer-probe sets: hs99999907_m1 (beta-2 microglobulin) and hs00265497_m1 (ribosomal protein L30). This combination was validated using pools of cDNA amplified in the TaqMan Human Endogenous Control Plate (Applied Biosystems). Selection of the two endogenous control genes was based on low cycle threshold (Ct) values that were not different among the cDNA reference pools. For each target, amplification was performed in duplicate reactions in a two-step cycle (denaturation, 95 °C for 15 s, annealing/extension at 62 °C for 60 s) for 40 cycles. Data output was analyzed using StepOne Software (v2.1, Applied Biosystems). To normalize target RNA expression, the geometric mean of the endogenous control RNAs was used to calculate ΔCt (Ct of the target - Ct geometric mean of endogenous controls). A ΔCt of 0 reflects equivalent expression between target and control. Differential mRNA abundance was quantified using the ΔΔCt method (ΔCt of individual sample - ΔCt of the same target in the reference pool).

### S2 Supplemental Figures and Legends

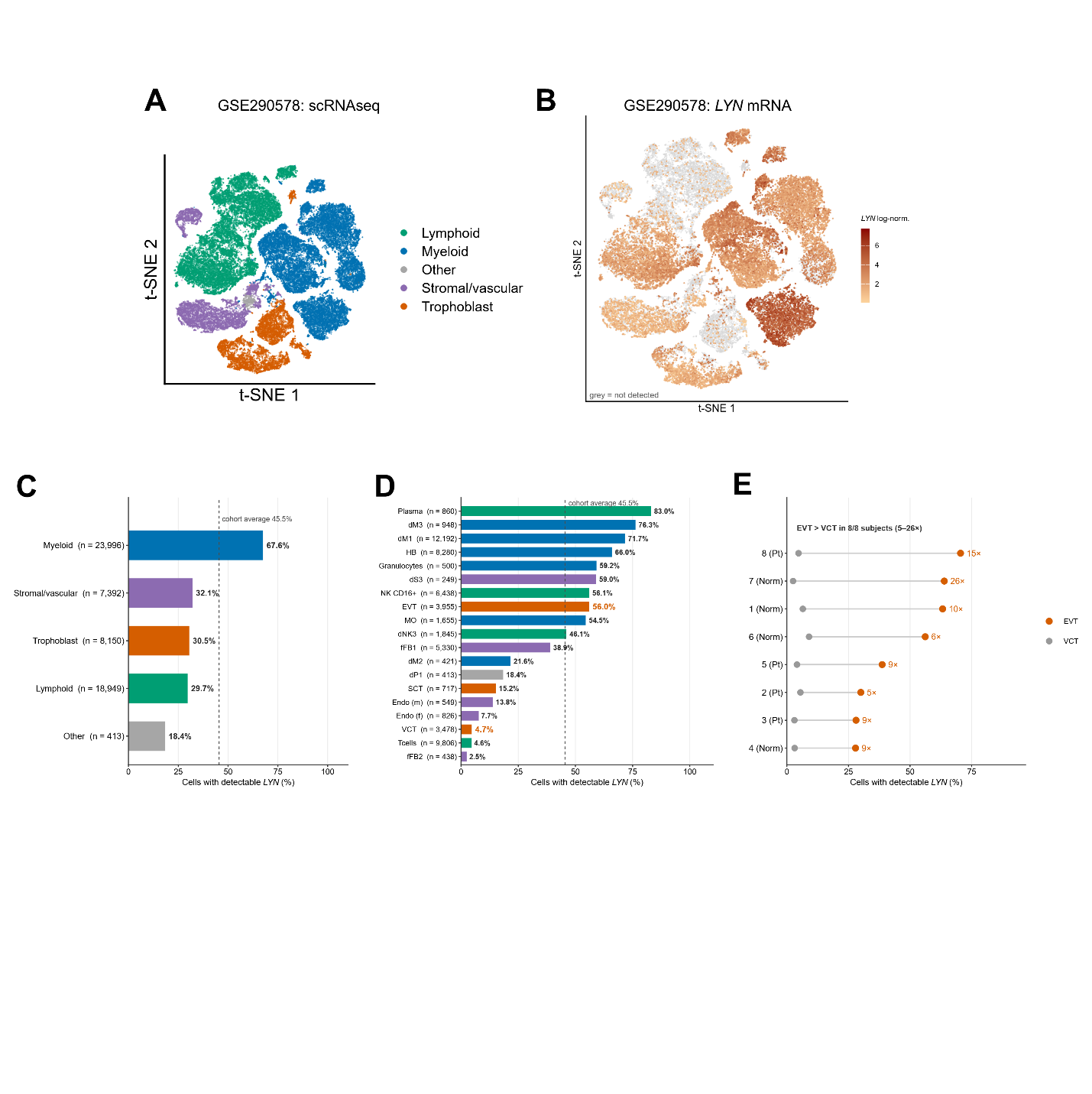

### Supplemental Fig. 1. *LYN* mRNA detection in individual placenta cell populations in the GSE290578 dataset. (A) Reanalysis of 58,900 individual placental cells from the GSE290578 dataset, visualized with t-distributed stochastic neighbor embedding (t-SNE) and color coded by predominant cell type per cluster. (B) *LYN* expression in the t-SNE plot as indicated by orange color saturation; 26,777 (45.5%) of placental cells had detectable *LYN*. Units are log_2_-transformed, scran-pooled-size-factor-normalized counts (offset by 1); size factors are centered at unity. (C) Percentages of cells per class with detectable *LYN*. (D) Percentages of stratified by detailed lineage group with detectable *LYN*. (E) Dumbbell plot showing *LYN* enrichment per subject comparing EVT (orange) versus VCT (gray) in the absence (Norm) or presence (Pt) of preeclampsia. Abbreviations: dM1-3, decidual macrophage subsets 1-3; dNK3, decidual natural killer cell subset 3; dS3, decidual stromal cell subset 3; dP1, decidual perivascular cell subset 1; Endo (f), fetal endothelial cell; Endo (m), maternal endothelial cell; EVT, extravillous trophoblast; fFB1-2, fetal fibroblast subsets 1-2; HB, Hofbauer cell (fetal placental macrophage); MO, monocyte; NK CD16+, CD16-positive natural killer cell; SCT, syncytiotrophoblast; VCT, villous cytotrophoblast.

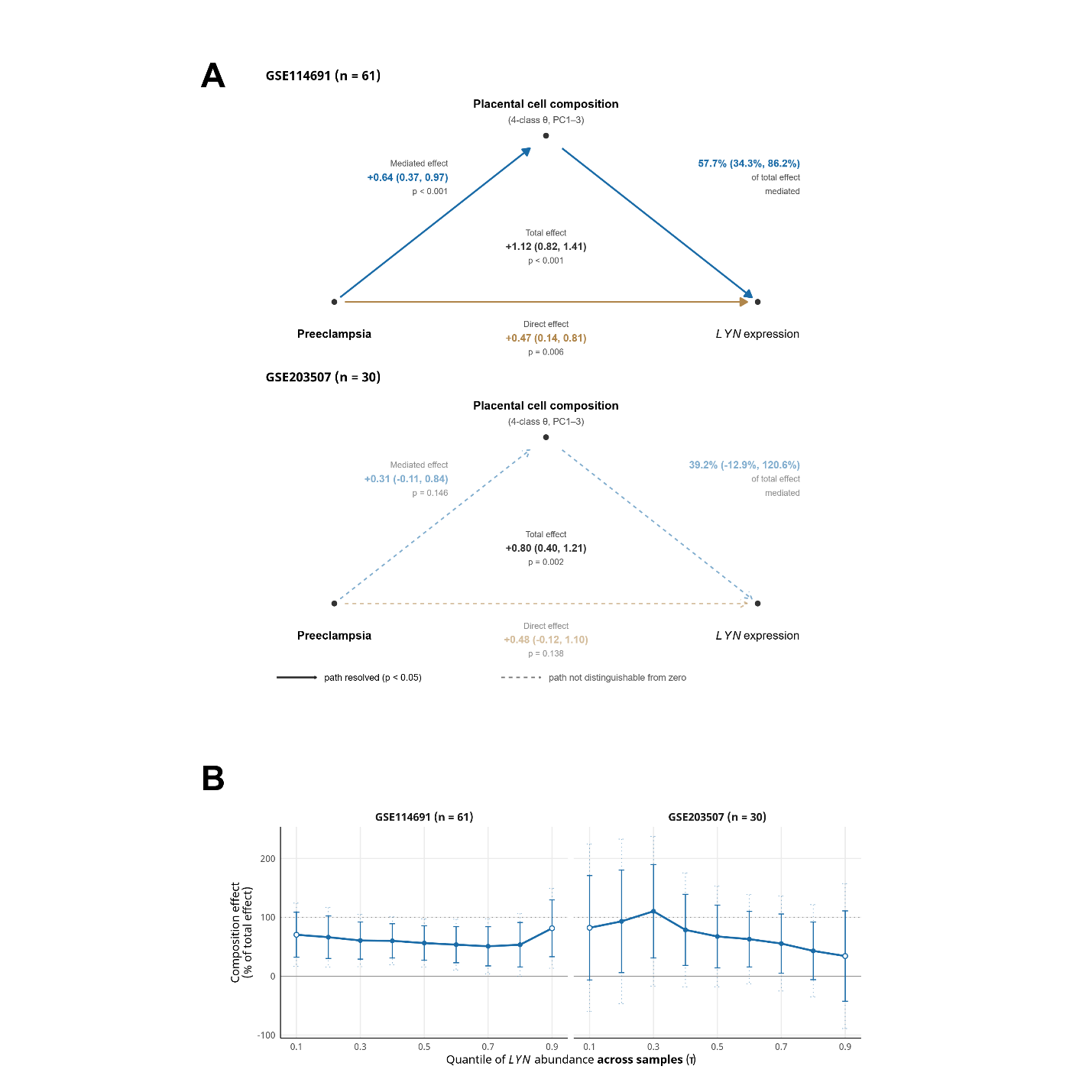

### Supplemental Fig. 2. Contribution of placental tissue composition to preeclampsia-associated *LYN* transcript abundance. (A) Causal mediation path diagrams showing g-formula causal mediation performed with CMAverse (v0.1.0; direct counterfactual imputation, 1,000 bootstrap resamples) for GSE114691 (top) and GSE203507 (bottom). In these models, the exposure was preeclampsia (PE), the outcome was variance-stabilized (vst) *LYN* transcript expression, and the mediator was the joint 3-principal-component representation of the 4-class deconvolved composition (trophoblast/stromal/vascular/immune). Solid arrows with closed heads denote paths resolved at p<0.05; dashed arrows with open heads indicate paths not distinguishable from zero. (B) Distributional decomposition (quantile-regression method) was performed using Counterfactual (v1.2); the composition effect has been expressed as a percentage of the total PE-associated *LYN* shift, estimated at each decile (τ=0.1-0.9) of *LYN* abundance across samples. Open markers denote the less reliable tail quantiles; solid error bars depict the derived bootstrap confidence intervals (CIs) on the composition effect, divided by the point estimate of the total effect; dotted error bars show the uniform (applicable to entire curve) bootstrap CIs.

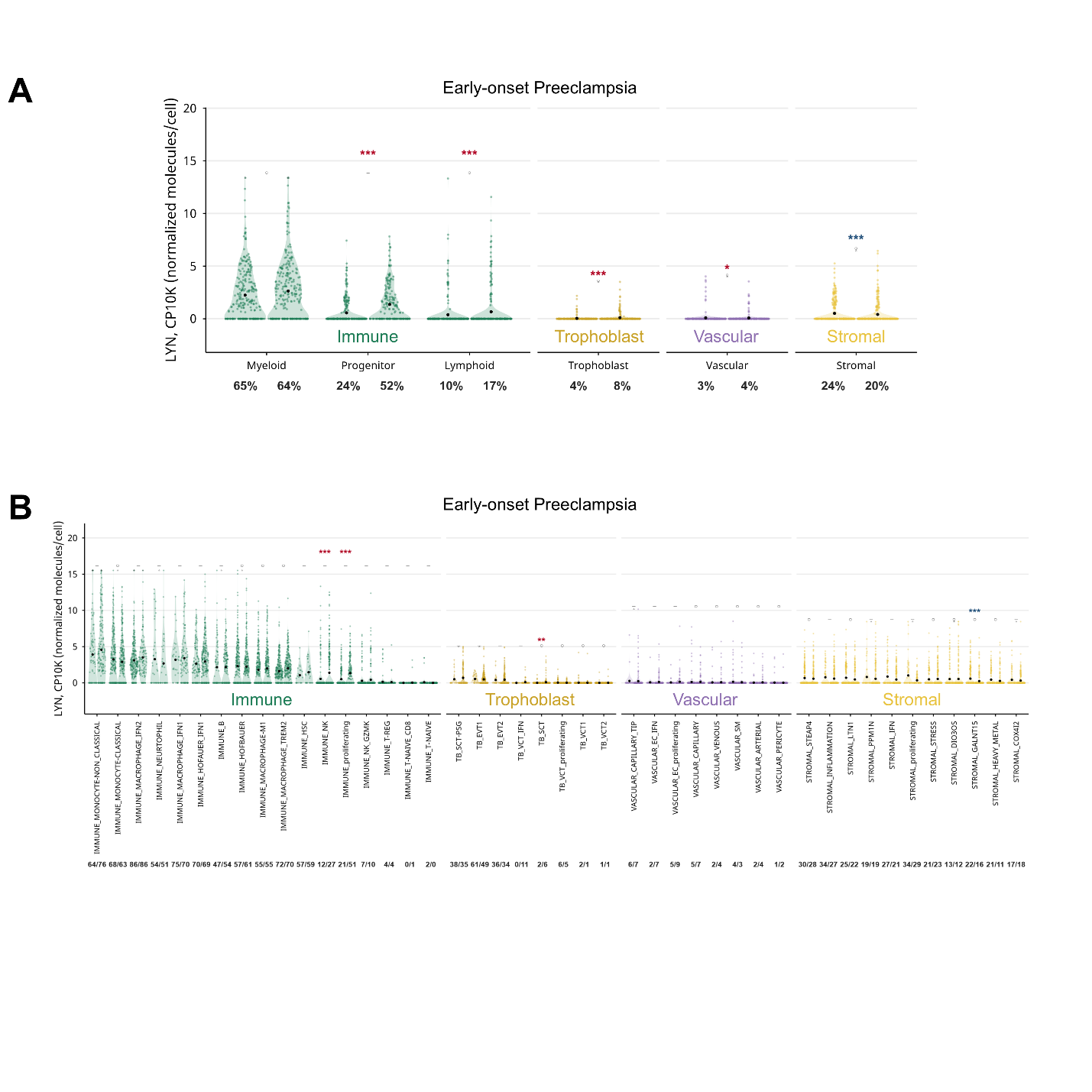

### Supplemental Fig. 3. Detailed stratification of lineage-specific *LYN* transcriptional expression in early-onset preeclampsia. (A) Sina plots showing normalized *LYN* counts (per 10,000 transcripts; CP10K) by lineage class for early-onset preeclampsia (PE) cases in the Admati atlas. Percentages under the axis are the detection rate (percent of cells with ≥1 detected *LYN* transcript). (B) Sina plots as in panel A, stratified by detailed cell class within each lineage category. Colored asterisks (red, increased in PE; blue, decreased in PE) represent two-sided Wilcoxon rank-sum tests on log(1 + CP10K), adjusted using the Benjamini-Hochberg FDR procedure. Black dots indicate mean values.
